# Neuropsychological and social predictors of participation in a deep brain stimulation study of Parkinson’s disease and dystonia

**DOI:** 10.1101/2024.05.29.24308133

**Authors:** Amelia Hahn, Ann A. Lazar, Stephanie Cernera, Simon Little, Sarah S. Wang, Philip A. Starr, Caroline A. Racine

## Abstract

**Objectives:** Participation is essential to DBS research, yet circumstances that affect diverse participation remain unclear. Here we evaluate factors impacting participation in an adaptive DBS study of Parkinson’s disease (PD) and dystonia.

**Methods:** Twenty participants were implanted with a sensing-enabled DBS device (Medtronic Summit RC+S) that allows neural data streaming in naturalistic settings and encouraged to stream as much as possible for the first five months after surgery. Using standardized baseline data obtained through neuropsychological evaluation, we compared neuropsychological and social variables to streaming hours.

**Results:** Marital status and irritability significantly impacted streaming hours (estimate=136.7, bootstrapped (^*b*^) *CI*^*b*^=45.0 to 249.0, *p*^*b*^=0.016, and estimate=-95.1, *CI*^*b*^=-159.9 to -49.2, *p*^*b*^=0.027, respectively). These variables remained significant after multivariable analysis. Composite scores on verbal memory evaluations predicted the number of hours of data streamed (*R*^*2*^=0.284, estimate=67.7, *CI*^*b*^=20.1 to 119.9, *p*^*b*^=0.019).

**Discussion:** Verbal memory impairment, irritability, and lack of a caregiver may be associated with decreased participation. Further study of factors that impact research participation is critical to the sustained inclusion of diverse participants.

## 1. Introduction

Participant recruitment and retention are among the most crucial and most challenging aspects of deep brain stimulation (DBS) clinical research [1,2], yet barriers to active participation remain understudied. Failure to retain participants can lead to inefficient use of resources, minimal research output, reduced statistical power to detect clinically important differences, participation bias, and delay of widespread access to novel treatment [3,4]. Factors associated with decreased participant involvement are complex and warrant thorough consideration to optimize recruitment and retention in not only studies employing DBS, but all investigations involving human participants **(Figure 1)**.

**Figure 1.**
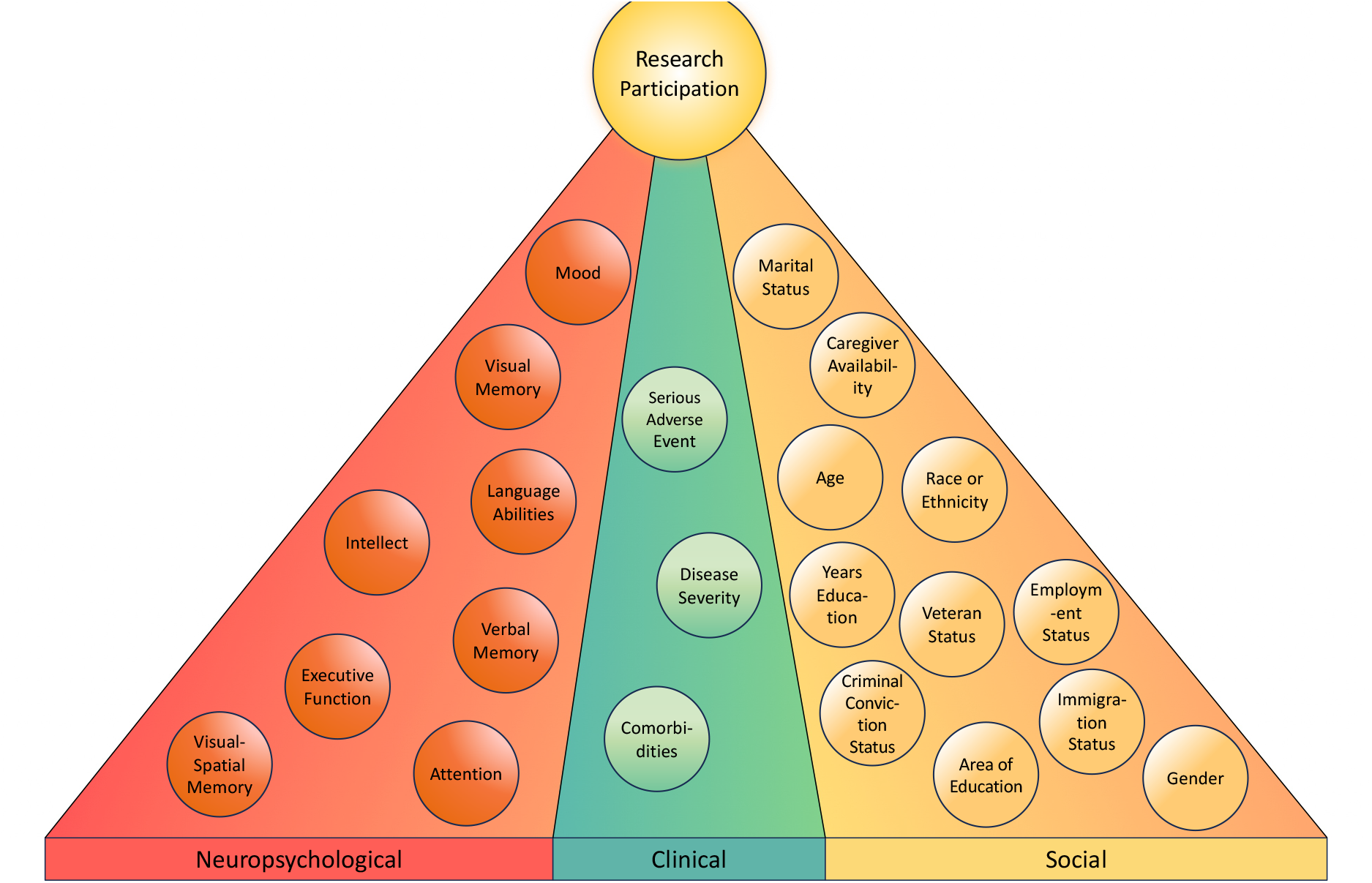
Neuropsychological, clinical, and social factors involved in the ability and willingness of an individual to contribute to time intensive clinical research. There are many variables to consider when evaluating an individual’s ability to participate, an understanding of which can inform strategies to maximize retention and long-term involvement of participants. Neuropsychological factors may impact an individual’s ability to learn data collection methods and juggle multiple research tasks. Clinical health factors may impact the time and physical extent to which a participant can contribute. Social factors may impact the willingness and ability of individuals to devote time to research.

Social factors can impact an individual’s willingness and ability to participate in research [5,6]. Social barriers to participation include race and ethnicity [2,7–9], age [9,10], lack of caregiver support [11], socioeconomic status [12], and language barriers [13], among others. These barriers particularly apply to DBS research for individuals with movement disorders. Access to standard of care DBS is not universal. Women, non-white individuals, and those of low socioeconomic status receive DBS as a treatment for movement disorders at a rate under-representative of the general patient population [14–17]. While studies have indicated unequal access to standard-of-care DBS, few have examined access to investigational DBS therapies, such as adaptive DBS. Concerns have been raised regarding long-term access to investigational brain implants following study completion as the cost of treatment increases [18], further emphasizing the importance of accounting for barriers to study inclusion prior to recruitment. Since research participants in DBS trials are often selected from individuals evaluated for standard-of-care DBS, additional social barriers to investigational DBS treatments likely exist and are not well understood. It is also unclear how social barriers in the recruitment process may impact retention, and further investigation could elucidate social factors that impact long-term, active participation in DBS studies.

In addition to social factors, neuropsychological factors may impact research participation. Individuals with severe cognitive impairment are typically withheld from standard-of-care DBS implantation due to concerns regarding poor clinical outcomes following implantation [19]. There may be neuropsychological factors identifiable during recruitment that could limit an individual’s ability to participate throughout the duration of a study. As innovations in the field increase the complexity and time intensity of DBS studies [20–22], understanding neuropsychological factors that may decrease participation will be instrumental in bolstering long-term retention of participants across a range of cognitive abilities.

While prior work has identified barriers and potential strategies to optimize the recruitment process in clinical and translational research [5,8,9,23], investigations of factors that may decrease long-term participation in time intensive DBS studies are unclear. The purpose of this study is (a) to identify social and neuropsychological factors impacting the number of hours of participation in a time intensive DBS study of twenty individuals with Parkinson’s disease (PD) or dystonia and (b) to determine methods of optimizing participation based on identified predictors.

## 2. Methods

### 2.1 Participation outcome: neural data streamed

Twenty individuals with PD or dystonia were enrolled in a study employing a sensing-enabled DBS device (Medtronic Summit RC+S) which allows concurrent recording of neural data in naturalistic settings while delivering therapeutic stimulation **(Table 1)**. Participants received either bilateral (two implantable pulse generators, *n*_*1*_=17) or unilateral (one implantable pulse generator, *n*_*2*_=3) RC+S implants. In our study, most participants were implanted with two electrodes per hemisphere, which required two Medtronic Summit RC+S implantable pulse generators. The sample size (*N*=20) was chosen based on the number of devices supplied by the manufacturer (40). To stream data, individuals place a communicator over the implantable pulse generator (IPG). The relay device connects via Bluetooth to custom research software on a Windows tablet and must remain resting over the implant for the duration of streaming (usually worn around the neck with a custom-designed sling). Participants must charge the tablet and communicator before streaming. Participants were encouraged to record as many hours of neural data as possible for the first five months following DBS implantation [21]. To qualitatively assess participant experiences with neural streaming, participants were sent the following survey questions via email:

**Table 1.**
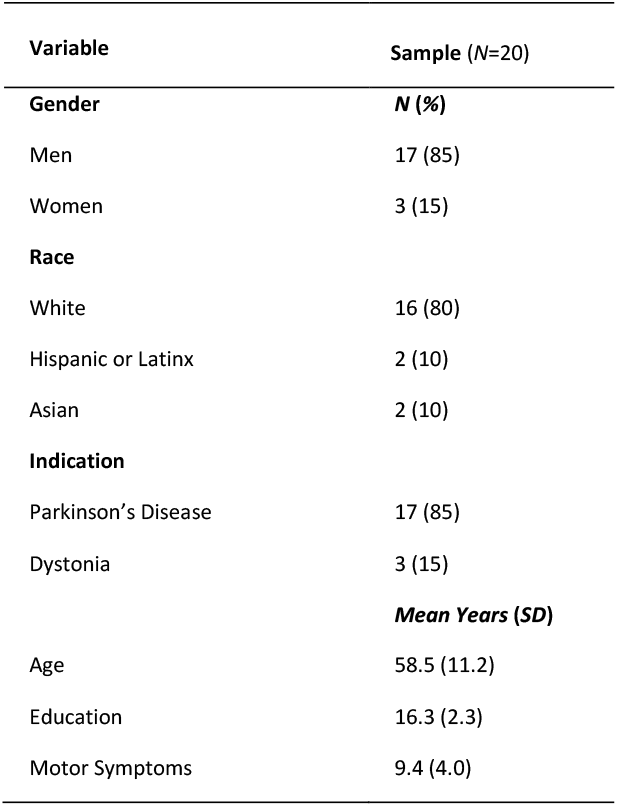
Participant demographics.

A. What does the process of streaming RC+S neural data look like for you?
B. What factors help you successfully stream data for a project?
C. What factors hinder your ability to successfully stream data for a project?
D. Do you have concerns about privacy and security when sensing?

### 2.2 Social and neuropsychological data obtained via clinical assessment

Clinical, psychological, and social variables were identified via clinic notes as well as interviews with individual participants and caregivers **(Supplementary Table 1a)**. All individuals underwent a baseline evaluation with the same clinical neuropsychologist (C. A. R.) prior to study enrollment. The neuropsychologist examined global cognition, estimated premorbid intellect, processing speed, verbal memory, visual and visio-spatial memory, language, and executive function, which are standard components of a typical neuropsychological assessment of DBS eligibility [24]. Neuropsychological variables were identified from assessments and standardized using available normative data that corrects for age and education **(Supplementary Table 1b)**.

### 2.3 Statistical analysis

The number of hours of data streamed in the first five months after lead implantation surgery were calculated using a custom MathWorks MATLAB® script. Clinical, psychological, and social variables were analyzed in relation to streaming hours using *t*-tests with a bias-corrected and accelerated bootstrapping (BCa) resampling technique to account for variability. Multivariable linear regression analyses were performed to adjust for statistically significant *t*-test results (two-sided *p*<0.05). Individual and composite *z*-scores from neuropsychological exams were analyzed in relation to participation hours using linear regression. We evaluated the form of the covariate by assessing non-linear terms (i.e., quadratic and cubic terms) for numeric variables. Since the non-linear terms were not statistically significant (two-sided *p*>0.05), we fit the linear term. Adjusted estimates and 95% BCa confidence intervals (CIs) were presented. Two-sided *p*-values <0.05 were considered statistically significant. All statistical analyses were run using IBM® SPSS® Statistics Version 29.0.0.0 (241).

### 2.4 Ethical compliance statement

This work was conducted as a sub-study of an adaptive DBS study which was approved by the institutional review board of University of California, San Francisco (IRB 18-24454). This study included all participants involved in the parent study. Written informed consent has been obtained from the participants.

## 3 Results

### 3.1 Caregiver relationship and participant mood impacts participation hours

No statistical significance was found in relation to participation hours in 50 of the 61 variables tested (82%). Married participants (*n*_*1*_=17, 85%) streamed more hours of data than those unmarried (*n*_*2*_=3, 15%, magnitude of mean difference [*μd*]=-127.9, *CI*^*b*^=-179.2 to -78.2, *p*^*b*^=0.003) **(Figure 2a)**. Participants whose caregiver attended the evaluation (*n*_*1*_=13, 65%), on average, streamed more hours than those whose caregiver did not attend (*n*_*2*_=7, 35%, *μd*=-81.9, *CI*^*b*^=-145.8 to -9.0, *p*^*b*^=0.042) **(Figure 2b)**. Individuals and spouses were questioned about the presence of increased irritability following their diagnosis during the clinical interview and ultimately a binary decision was made using the neuropsychologist’s judgment. Individuals or their spouse who indicated increased irritability or agitation since their diagnosis (*n*_*1*_=9, 45%) streamed fewer hours of data than the group that did not indicate increased irritability (*n*_*2*_=11, 55%, *μd*=86.4, *CI*^*b*^=15.7 to 156.3, *p*^*b*^=0.036) **(Figure 2c)**. Within the group of married participants, the irritable group (*n*_*1*_=8, 47%) streamed fewer hours of data than the group that was not irritable (*n*_*2*_=9, 53%, *μd*=114.9, *CI*^*b*^=46.7 to 178.8, *p*^*b*^=0.014) **(Figure 2d)**. After adjusting for marital status, caregiver attendance, and irritability in a linear regression model of streaming hours in the first five months of the study, both marital status and irritability remained statistically significant (*p*^*b*^=0.012 and *p*^*b*^=0.019, respectively), but caregiver attendance did not (*p*^*b*^=0.843, **Figure 2e**).

**Figure 2.**
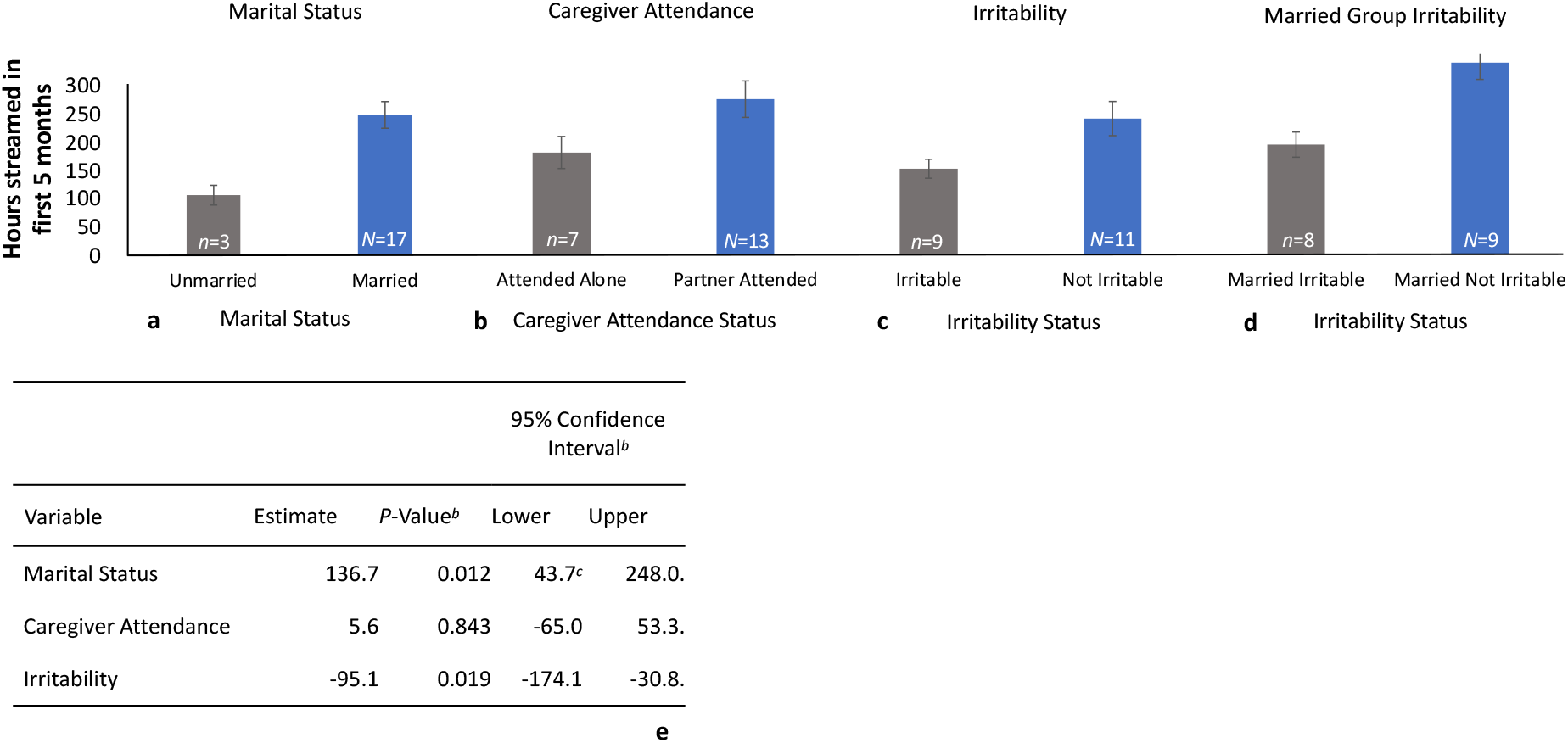
Impact of the caregiver relationship and participant changes in mood on the number of data hours streamed. Difference in mean data hours streamed between **(a)** married and unmarried participants (magnitude of mean difference [*μd*]=-127.9, *CI*^*b*^=-179.2 to -78.2, *p*^*b*^=0.003), **(b)** individuals whose caregiver attended the neuropsychological exam and those who attended alone (*μd*=-81.9, *CI*^*b*^=-145.8 to -9.0, *p*^*b*^=0.042), **(c)** the irritable and not-irritable group (*μd*=86.4, *CI*^*b*^=15.7 to 156.3, *p*^*b*^=0.036), and **(d)** the irritable and not-irritable group within the cohort of married participants (*μd*=114.9, *CI*^*b*^=46.7 to 178.8, *p*^*b*^=0.014). When questioned in the clinical interview about the presence of increased irritability following their diagnosis, a binary decision for inclusion in the irritable vs not-irritable groups was made using the neuropsychologist’s judgment. **(e)** Multivariable analysis shows marital status and irritability findings remain significant after adjustment. After adjusting for marital status, caregiver attendance, and irritability in a linear regression model of participation hours in the first five months of the study (outcome), both marital status and irritability remain statistically significant (*p*^*b*^=0.012 and *p*^*b*^=0.019 respectively), but caregiver attendance does not (*p*^*b*^=0.843). ^*b*^ *= bias corrected and accelerated (BCa) bootstrap value* ^*c*^ *= percentile method value*

### 3.2 Verbal memory scores predict number of hours of participation

Regression analysis showed a positive association between streaming hours and *z*-scores of list recall after a 3-minute delay (*R*^2^=0.276, estimate=68.0, *CI*^*b*^=16.6 to 113.0, *p*^*b*^=0.014) **(Figure 3a)** and a 12-minute delay (*R*^2^=0.236, estimate=56.4, *CI*^*b*^=8.3-105.4, *p*^*b*^=0.026) **(Figure 3b)**. Similarly, we found an association between streaming hours and both immediate (*R*^2^=0.307, estimate=42.3, *CI*^*b*^=11.4 to 74.5, *p*^*b*^=0.034) **(Figure 3c)** and 10-minute delayed (*R*^2^=0.218, estimate=44.4, *CI*^*b*^=6.5 to 73.3, *p*^*b*^=0.024) **(Figure 3d)** story recall. A composite *z*-score across all verbal memory variables also significantly predicted participation hours (*R*^*2*^=0.284, estimate=67.7, *CI*^*b*^=20.1 to 119.9, *p*^*b*^=0.019) **(Figure 3e)**.

**Figure 3.**
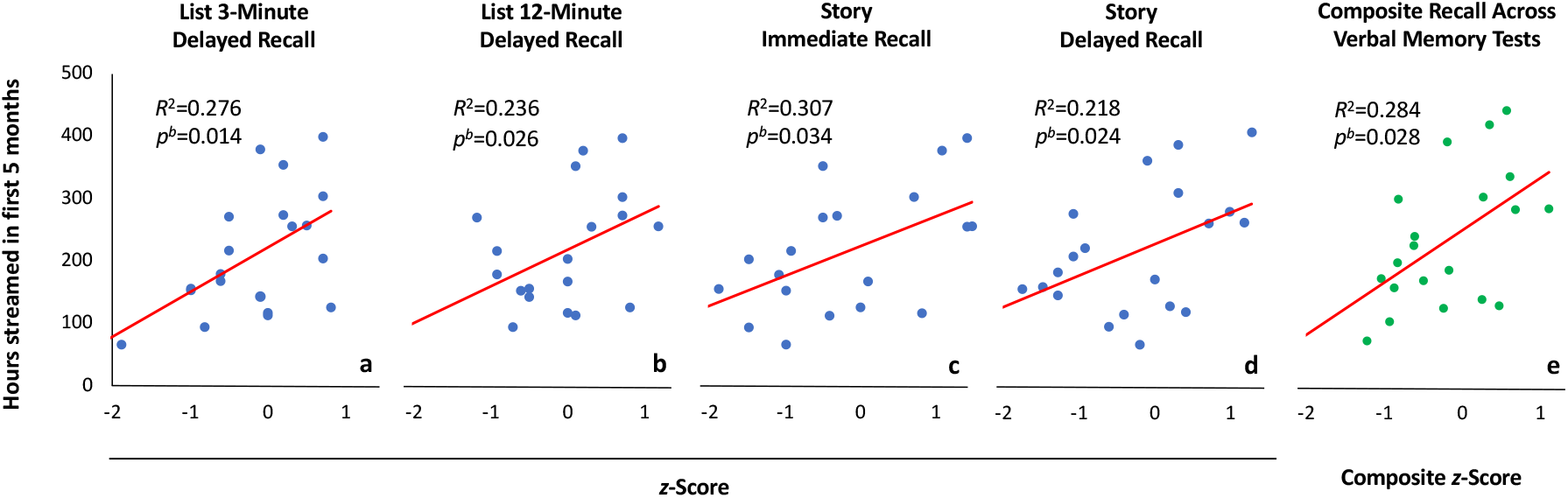
Verbal memory scores predict the number of hours of participation. Significant positive correlations between participation hours and *z*-scores of **(a)** list recall after a 3-minute delay, **(b)** list recall after a 12-minute delay, **(c)** immediate story recall, and **(d)** story recall after a 10-minute delay. **(e)** A significant positive correlation was found between participation and memory for composite *z*-scores across all verbal memory exams.

### 3.3 Qualitative responses to participant surveys

Four of our highest-contributing participants responded to the survey. They noted communication from the research team (i.e., clear instruction and reminders) as helpful in successfully streaming data, while lack of communication hindered streaming. Technological difficulties interfered with streaming (i.e., internet connectivity and research software issues). One participant suggested that forgetting steps required to operate research equipment led to incomplete or failed data collection, and another said phone reminders and notes facilitated successful streaming. Participants did not indicate privacy concerns regarding streaming (as one stated, “I don’t think my brain data is something that anyone other than researchers can understand or use”). This limited sample may not fully reflect the views of our entire cohort but does help us more fully understand the experience of our most active participants and suggests topics for future exploration.

## 4. Discussion

In this complex DBS study requiring at-home data streaming, verbal memory impairment, irritability, marital status, and caregiver attendance at the neuropsychological evaluation impacted participation as measured by the number of hours of data streamed at home. Marital status emerged as the most significant social predictor of participation, while verbal memory was a significant neuropsychological predictor. Aside from marital status, caregiver attendance, participant irritability, and verbal memory, no significant trends were found for other variables tested. There were no relationships shown between the number of streaming hours and age, gender, race or ethnicity, clinical diagnosis, years of education, employment, immigration, or criminal conviction status. While increased irritability following the diagnosis of a movement disorder decreased streaming hours, there were no trends found for depression, anxiety, apathy, or impulsivity. Across neuropsychological variables, estimated premorbid intellect, processing speed, attention, visual memory, language, and executive function did not impact streaming hours. Additionally, while not quantitatively assessed in this study, ethics are an important consideration in sensing trials and may impact participation. This is a crucial area for future studies on motivation to participate in DBS trials, with prior studies indicating that concerns about privacy and security can affect the decision to enroll in a brain sensing study [25].

The widespread trend across four different measures of verbal memory may indicate that verbal memory deficits hindered participants’ ability to remember when and how to stream data. Although participants were instructed on the use of the data streaming interface at the start of the study, those with memory impairment may have forgotten prior instruction, had difficulty recalling troubleshooting procedures, and/or forgotten to stream during their daily activities. This is consistent with prior research suggesting that cognitive decline can negatively impact research participation [26]. The difficulties participants experienced in remembering to stream could be understandable given the inherent complexity of the RC+S data collection user interface and the learning required to operate the devices independently at home **(Section 2.1)**, which is supported by survey responses. Similar factors have been shown to affect the decision to enroll in sensing-enabled neural interfaces, which have identified complicated DBS technology as a deterrent to enrollment [25]. Trends in verbal memory were driven primarily by delayed recall.

Participants who did not have a spouse or partner may have received less assistance operating study-related devices, making it more difficult to learn and execute the steps required to ensure successful data collection. This is consistent with previous findings indicating that caregivers bolster the success of a clinical trial [27,28]. Previous research on motivation to participate in DBS studies has identified motivated caregivers as being influential to the successful enrollment of a participant, and our findings offer corroborative evidence that a caregiver may increase motivation to continue participating beyond enrollment [25]. It is unclear how increased irritability may have decreased participation hours, in part because the irritability assessment consisted of a binary choice lacking a grading scale. Authors speculate that individuals who were more irritable may have abandoned data collection tasks out of agitation or had trouble collaborating with others to facilitate successful streaming. The significantly fewer hours contributed by married participants who also indicated increased irritability may suggest difficulty collaborating with a partner, which authors have noted in some participants. Partner assistance may have had an additional benefit for participants with memory impairment, with a partner or spouse improving recall of study directions. This is consistent with findings related to the Joint Memory Effect, which involves enhancement of memory when collaborating with a partner [29].

### 4.1 Strategies for bolstering retention

While we have identified factors that may decrease research participation, this does not warrant the exclusion of individuals from research based on these factors. Although they participated less overall, these individuals still contributed considerable hours to the study. Rather than serving as a basis for participant exclusion, the identification of these factors during the recruitment period can serve to inform strategies for individualized support to maximize their contributions.

The factors identified in this study inform retention strategies in complex, time intensive DBS and non-DBS studies. The verbal memory trends as well as qualitative survey responses underscore the necessity of effective communication between research staff and participants. Supplementing verbal instructions with written materials and follow-up reminders may help ensure participants complete tasks as directed. Scheduled, consistent communication from a designated member of the research team may encourage compliance, particularly for long-term monitoring of participants. For participants without a caregiver or task partner assisting with research responsibilities, frequent reminders over text and email as well as live instruction via phone call, video chat, and in-person communication may significantly increase the quantity of data individuals are able to contribute. Visiting participants in their homes to assist with data streaming in a naturalistic setting, which was necessary for this study, can be particularly effective for individuals lacking support at home. These strategies are consistent with previous studies showing that individualized support based on the unique needs of each participant can increase their willingness and ability to contribute to research [30].

### 4.2 Limitations

The small sample size of this study, relative lack of racial or gender diversity, and small sample of unmarried participants limit the generalizability of these results. Variability was introduced to this data set using bias-controlled and accelerated bootstrapping for all *t*-tests, and Mann-Whitney-Wilcox tests yielded the same results of significance. Retrospective data collection and analysis may limit findings. Several variables identified through medical records and clinical interview were subject to clinician’s judgment and were not measured with a standardized questionnaire. The results of this study should be interpreted with caution until confirmatory studies can be performed due to the small sample size and large number of variables tested.

### 4.3 Conclusions

Verbal memory impairment, increased irritability, and lack of a partner reduced participation in this time intensive DBS study. Based on this study, strategies to optimize participation include frequent contact with the research team, instruction with both verbal and visual details, text or call reminders before study visits or tasks, and additional guidance for those without a caregiver. Further understanding of factors contributing to research participation is critical to the development of protocols that allow for the successful recruitment, engagement, and retention of a diverse set of individuals.

## Data Availability

All data produced in the present study are available upon reasonable request to the authors

## Acknowledgements

We thank B. Swinnen and E. Ubeda-Matzilevich for a critical review of this manuscript and W. Chiong for neuroethical oversight of this study.

## Highlights

- Social and neuropsychological factors impact participation in DBS studies
- Memory impairment, lack of a caregiver, and irritability reduce participation
- Complex DBS studies would benefit from individualized participant support strategies

## Figure Legends

**Supplementary Table 1.**
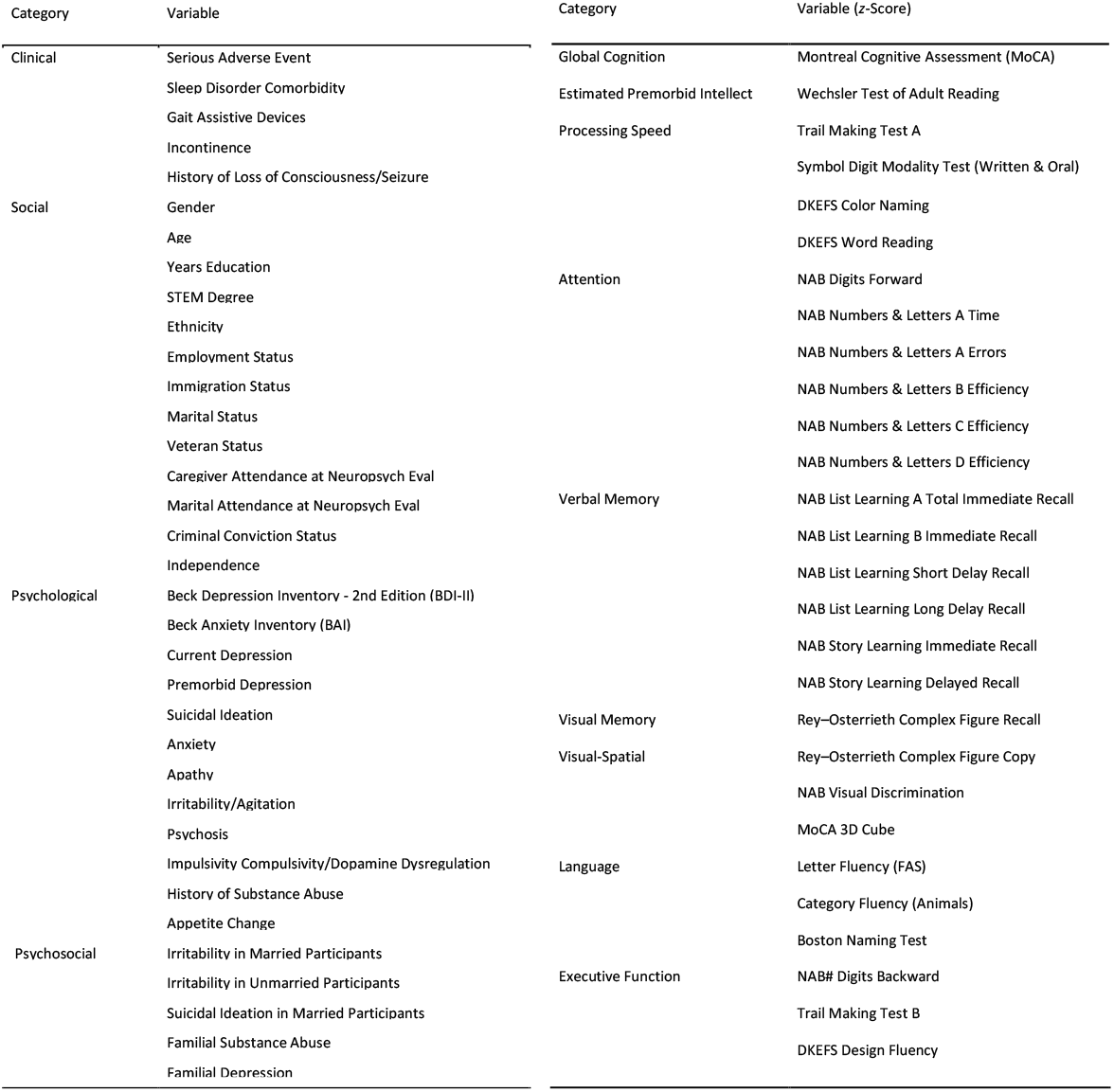
Variables tested. Variables were identified via **(a)** clinic and interview notes and **(b)** neuropsychological testing. NAB = Neuropsychological Assessment Battery, DKEFS = Delis-Kaplan Executive Function System.

## Notes

**Sources of Financial Support:** This work was a sub-study of an adaptive deep brain stimulation study supported by National Institute of Health awards NINDS UH3NS100544, R01NS131405-01, and U24NS113667. This publication was also supported by the National Center for Advancing Translational Sciences, National Institutes of Health, through UCSF-CTSI Grant Number CTSI UL1 TR001872. The sponsor was not involved in this study’s design, the collection, analysis, or interpretation of data, nor in writing the report, nor the decision to submit the article for publication.

**Conflict of Interest Statement:** Nothing to declare.

### Competing Interest Statement

The authors have declared no competing interest.

### Funding Statement

This work was a sub-study of an adaptive deep brain stimulation study supported by National Institute of Health awards NINDS UH3NS100544, R01NS131405-01, and U24NS113667. This publication was also supported by the National Center for Advancing Translational Sciences, National Institutes of Health, through UCSF-CTSI Grant Number CTSI UL1 TR001872. The sponsor was not involved in this study's design, the collection, analysis, or interpretation of data, nor in writing the report, nor the decision to submit the article to medRxiv.

### Author Declarations

This work was conducted as a sub-study of an adaptive DBS study which was approved by the institutional review board of University of California, San Francisco (IRB 18-24454).

